# Application of simultaneous uncertainty quantification for image segmentation with probabilistic deep learning: Performance benchmarking of oropharyngeal cancer target delineation as a use-case

**DOI:** 10.1101/2023.02.20.23286188

**Authors:** Jaakko Sahlsten, Joel Jaskari, Kareem A. Wahid, Sara Ahmed, Enrico Glerean, Renjie He, Benjamin H. Kann, Antti Mäkitie, Clifton D. Fuller, Mohamed A. Naser, Kimmo Kaski

## Abstract

**Background:** Oropharyngeal cancer (OPC) is a widespread disease, with radiotherapy being a core treatment modality. Manual segmentation of the primary gross tumor volume (GTVp) is currently employed for OPC radiotherapy planning, but is subject to significant interobserver variability. Deep learning (DL) approaches have shown promise in automating GTVp segmentation, but comparative (auto)confidence metrics of these models predictions has not been well-explored. Quantifying instance-specific DL model uncertainty is crucial to improving clinician trust and facilitating broad clinical implementation. Therefore, in this study, probabilistic DL models for GTVp auto-segmentation were developed using large-scale PET/CT datasets, and various uncertainty auto-estimation methods were systematically investigated and benchmarked.

**Methods:** We utilized the publicly available 2021 HECKTOR Challenge training dataset with 224 co-registered PET/CT scans of OPC patients with corresponding GTVp segmentations as a development set. A separate set of 67 co-registered PET/CT scans of OPC patients with corresponding GTVp segmentations was used for external validation. Two approximate Bayesian deep learning methods, the MC Dropout Ensemble and Deep Ensemble, both with five submodels, were evaluated for GTVp segmentation and uncertainty performance. The segmentation performance was evaluated using the volumetric Dice similarity coefficient (DSC), mean surface distance (MSD), and Hausdorff distance at 95% (95HD). The uncertainty was evaluated using four measures from literature: coefficient of variation (CV), structure expected entropy, structure predictive entropy, and structure mutual information, and additionally with our novel *Dice-risk* measure. The utility of uncertainty information was evaluated with the accuracy of uncertainty-based segmentation performance prediction using the Accuracy vs Uncertainty (AvU) metric, and by examining the linear correlation between uncertainty estimates and DSC. In addition, batch-based and instance-based referral processes were examined, where the patients with high uncertainty were rejected from the set. In the batch referral process, the area under the referral curve with DSC (R-DSC AUC) was used for evaluation, whereas in the instance referral process, the DSC at various uncertainty thresholds were examined.

**Results:** Both models behaved similarly in terms of the segmentation performance and uncertainty estimation. Specifically, the MC Dropout Ensemble had 0.776 DSC, 1.703 mm MSD, and 5.385 mm 95HD. The Deep Ensemble had 0.767 DSC, 1.717 mm MSD, and 5.477 mm 95HD. The uncertainty measure with the highest DSC correlation was structure predictive entropy with correlation coefficients of 0.699 and 0.692 for the MC Dropout Ensemble and the Deep Ensemble, respectively. The highest AvU value was 0.866 for both models. The best performing uncertainty measure for both models was the CV which had R-DSC AUC of 0.783 and 0.782 for the MC Dropout Ensemble and Deep Ensemble, respectively. With referring patients based on uncertainty thresholds from 0.85 validation DSC for all uncertainty measures, on average the DSC improved from the full dataset by 4.7% and 5.0% while referring 21.8% and 22% patients for MC Dropout Ensemble and Deep Ensemble, respectively.

**Conclusion:** We found that many of the investigated methods provide overall similar but distinct utility in terms of predicting segmentation quality and referral performance. These findings are a critical first-step towards more widespread implementation of uncertainty quantification in OPC GTVp segmentation.

## Introduction

Oropharyngeal cancer (OPC), a type of head and neck squamous cell carcinoma (HNSCC), is a widespread and debilitating disease ^1^. A core treatment modality in OPC patient care is radiotherapy (RT). The current standard of care for OPC RT relies on manually generated segmentation of the primary gross tumor volume (GTVp) by clinical experts as a target structure to deliver RT dose. However, the GTVp in OPC is notorious for being one of the most difficult structures amongst all cancer types to perform accurate segmentation for RT planning due to its exceptionally high interobserver variability ^2–4^. Subsequently, GTVp segmentation has been cited as the single largest factor of uncertainty in RT planning ^5,6^. Therefore, automated approaches that can reduce interobserver variability are paramount to improving the current OPC RT workflow.

Deep learning (DL) has increasingly been used in the OPC RT space for automatically segmenting organs at risk ^7,8^ and target structures ^9–12^. Impressively even for GTVp segmentation, several DL approaches have boasted exceptionally high performance in terms of volumetric and surface-level agreement with the ground-truth segmentations ^13^. Importantly, the DL-based auto-segmentation has been shown to be superior to expected expert interobserver variability ^14^, thereby highlighting its potential utility as a support tool for accurate clinical decision making. Notably many of these advances in OPC GTVp segmentation have been spurred by open-source data challenges ^15^, namely the HEad and neCK TumOR (HECKTOR) PET/CT tumor segmentation challenge ^14,16^. However, while there exist a deluge of DL-based OPC auto-segmentation approaches that demonstrate potentially clinically acceptable performance in terms of geometric measures, the relative confidence (i.e., uncertainty) with which these models generate predictions remains a relatively unexplored domain.

Quantification of the DL model uncertainty is crucial to improve the trust of clinicians’ in model predictions and to facilitate the clinical implementation of these technologies ^17^. Within RT, segmentation is a clear and well-discussed application space for uncertainty estimation ^18^. This is particularly relevant for RT target structures (i.e., OPC GTVp) where high interobserver variability is expected. While the performance of OPC GTVp auto-segmentation models is seemingly impressive, the actual clinical utility for most of these methods has yet to be solidified due to a lack of investigations on model uncertainty. Previous work in DL uncertainty estimation has been extensively investigated in segmenting lung-related ^19–21^ and brain-related ^22–24^ structures. While DL uncertainty estimation has been applied to a broad range of HNSCC-related classification tasks ^25–28^ and dose prediction ^29^, only a limited number of studies have investigated uncertainty estimation for 3-dimensional HNSCC medical image segmentation, predominantly for nasopharyngeal cancer ^30^ or organs at risk ^31,32^; to our knowledge only one study has attempted to investigate segmentation uncertainty estimation in OPC ^33^. Therefore, there exists a significant gap in knowledge about how to construct DL auto-segmentation models that lend themselves to uncertainty estimation and subsequently how to quantify the model uncertainty at individual patient and voxel-wise levels for OPC GTVp segmentation.

In this study, we developed probabilistic DL GTVp auto-segmentation models using large-scale PET/CT datasets and systematically investigated various uncertainty estimation methods for voxel-wise and patient-level uncertainty. We employed several quantitative evaluation methods to link uncertainty measures to known performance measures and qualitatively investigated uncertainty results.

## Methods

In this section, we present the datasets we used for training and evaluation of our models, describe the DL models we employ, introduce the uncertainty metrics that are used to quantify the model uncertainty, and list all the performance evaluation metrics used for the experiments.

### Dataset

For this study, we utilized two main OPC patient datasets containing PET/CT data: 1.) the publicly available 2021 HECKTOR Challenge training dataset ^16^, and 2.) an external validation dataset from The University of Texas MD Anderson Cancer Center (MDA). The HECKTOR dataset contained 224 OPC patients with co-registered PET/CT scans with manually generated GTVp segmentation masks from multiple clinician annotators (one annotator per scan). Additional details on the HECKTOR dataset can be found in the corresponding overview paper ^16^. The MDA external validation dataset contained 67 OPC patients with co-registered PET/CT scans with manually generated GTVp segmentation masks from a single clinician annotator (S.A.). Additional details on the MDA external validation dataset can be found in **Appendix A**. The MDA external validation dataset was retrospectively collected under a HIPAA-compliant protocol approved by the MDA institutional review board (RCR03-0800) which gave ethical approval for this work.

For model training and evaluation all data was resampled into 1 mm isotropic pixel spacing, 1 mm slice thickness, and cropped into 144 × 144 × 144 voxel sized volumes centered around the GTVp segmentation. The CT scans were windowed at [-200, 200] Hounsfield Units to [-1, 1] range and the PET scans were z-score normalized. The models were trained using a 5-fold cross-validation scheme on the HECKTOR dataset. For the performance evaluation of the model, the MDA external validation dataset was used.

### Probabilistic deep learning models

Conventional DL approaches have been observed to be overconfident in the predictions they make, which means that the probability estimates they provide do not correspond to the observed likelihood of them being correct ^34^. The Bayesian approach has been described to show promise in improving uncertainty estimation and calibration of DL methods ^35^ We chose to investigate two approximations of Bayesian inference in DL, i.e. the Deep Ensemble and the Monte Carlo (MC) Dropout Ensemble that are two popular methods used in uncertainty-aware DL for medical segmentation tasks.

The DL architecture used for all the models was a 3D residual U-net from the Medical Open Network for AI (MONAI) ^36^, due to its established success in OPC GTVp segmentation ^10,12,16,37^. This architecture has two input channels for both modalities, i.e., the CT and PET, and a single output channel with the sigmoid activation function. The default MONAI Residual U-net model was used with an encoder consisting of five convolution blocks with 16, 32, 64, 128, and 256 channels, a decoder mirroring the channel count, and a feature concatenation from the decoder to the respective encoder block. Each block has two convolution layers each followed by instance normalization, dropout and parametric ReLU layers, and a residual connection with convolution between the block input and the last layer. The MC Dropout method applies the dropout stochastic regularization layer during test-time ^38^, whereas the output was deterministic with the Deep Ensemble.

Both the Deep Ensemble and the MC Dropout Ensemble consisted of five models that were each trained using 5-fold cross validation for the HECKTOR dataset. From a Bayesian point of view, the posterior predictive distribution of a Deep Ensemble is approximated with a uniform mixture of the individual networks in the ensemble, i.e., the average of the predictions of the individual networks is approximate Bayesian inference. With the MC Dropout Ensemble, the uniform mixture is over multiple networks with MC dropout, and the predictive distribution is approximated by the average over Monte Carlo samples from each of these networks ^39^. In practice, we used 60 MC samples from each of the five ensemble members for the approximation.

For both the Deep Ensemble and the MC Dropout Ensemble, we searched for optimal hyperparameters using the HECKTOR dataset. The hyperparameters we considered were the loss function, specifically the choice between the Dice loss and the sum of Dice and Binary Cross-Entropy losses, as well as the dropout rate, searched from the range of [0.1, 0.9]. The hyperparameter selection criterion was based on the quality of the uncertainty estimation with the area under the Dice similarity coefficient referral curve on the validation data (described more in Uncertainty measures).

### Uncertainty measures

The Bayesian DL methods provide probability estimates that should better correspond to the observed likelihood of correct predictions they make, i.e., better calibration. To quantify the uncertainty of these models, a widely used measure in the literature is the information theoretic entropy of the predictive distribution ^39–42^. It measures the spread of the probability mass, as it attains the maximum value for a uniform distribution over the possible events and the minimum when the mass is concentrated on a single class ^41^. However, recent works in uncertainty-aware medical segmentation have demonstrated that other measures of uncertainty have high correlation with the performance of the DL model ^20,43,44^.

Let *x* denote the input scans of the patient and *y* the target GTVp volume. In the binary segmentation task, the Deep Ensemble and the MC Dropout Ensemble produce samples of the predictive distribution; *p*^*(m)*^(*y*_*i,j,k*_ =1|*x*), where *m* is the index of the sample and *i,j,k* denote voxel coordinates. In the case of MC Dropout, Roy et al. ^43^ and Hoebel et al. ^20^, proposed three so-called structure-wise uncertainty measures; the coefficient of variation (CV), the mean pairwise Dice, and the structure expected entropy. We consider the segmentation of GTVp, i.e., a single structure, and thus the structure-wise uncertainty is only computed for the positive class. The CV is defined as:

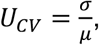

where *µ* and *σ* are computed as follows:

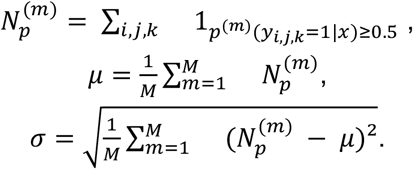

The CV thus measures the variation of the volume of a segmentation observed in the samples.

The structure expected entropy, using the definition in Hoebel et al., is as follows:

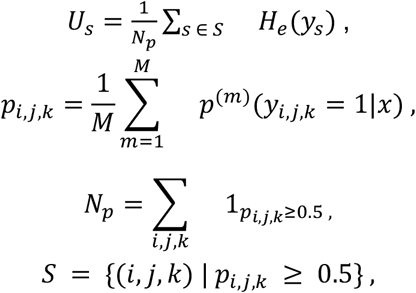

where *H*_*e*_ is the so-called *expected entropy*:

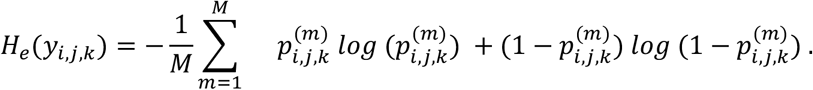

Thus the structure expected entropy measures the average of the expected entropy over the voxels that were considered as positive based on the average of the Monte Carlo samples. In Mukhoti and Gal ^45^, two other entropy-based uncertainty measures were used. Namely, predictive entropy and mutual information. As the expected entropy is a part of the mutual information, we also experimented by utilizing the predictive entropy:

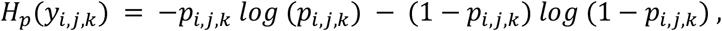

and the mutual information:

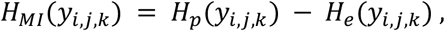

when calculating the structure entropy, denoted as *U*_*p*_ and *U*_*MI*_. These three types of entropy-based uncertainty measures have been described to account for the total uncertainty in the case of predictive entropy, the aleatoric uncertainty in the case of the expected entropy, and the epistemic uncertainty in the case of mutual information ^46^.

The mean pairwise Dice used in Roy et al. and Hoebel et al. is computed as the average Dice coefficient of all the pairs constructed from the samples. In order to compute the measure, we need *M*(*M*−1)/2 comparisons, which is 44850 comparisons for the MC Dropout Ensemble; we deemed the measure prohibitive to compute in practice. Inspired by the fusion of the Dice coefficient and the uncertainty estimation, we developed a novel uncertainty measure that we call *Dice-risk*, which is based on the expected conditional risk introduced in a recent work ^47^. In this work, the authors noted that the entropy-based uncertainty measures can be interpreted as computing the average negative log-likelihood provided that the target is distributed as the network predictive distribution describes. They showed that by replacing the negative log-likelihood with some other loss function, novel uncertainty measures could be derived. We utilized the Dice-loss, defined as 1 − *DSC*, as the loss function.

The Dice-risk uncertainty measure is defined as follows:

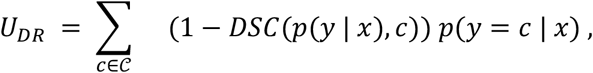

where *𝒞* is the set of all possible segmentations. Since the expectation cannot be computed in practice, we utilize a stochastic estimate of the Dice-risk, by computing the expectation with Monte Carlo approximation:

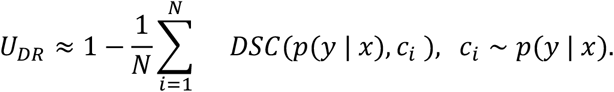

It is thus estimated in a “doubly stochastic” manner, since *p*(*y* | *x*) is also estimated with Monte Carlo approximation.

### Performance evaluation

We evaluated the segmentation performance with the Dice similarity coefficient (DSC), mean surface distance (MSD), and mean Hausdorff distance at 95% (95HD), specifically, the median value of these metrics. These metrics were selected because of their ubiquity in literature and ability to capture both volumetric overlap and boundary distances ^48,49^. For the 5-fold cross validation results, we report the mean and standard error of the mean (SEM) of the metrics computed on each fold, whereas for the holdout set we report the point estimates. The model output was resampled into original resolution with nearest-neighbor sampling and evaluated against original resolution segmentations. The performance of MSD and 95HD was evaluated in millimeters. When comparing the segmentation model metrics, we implemented Wilcoxon signed rank tests with p-values less than or equal to 0.05 considered as significant. Statistical comparisons were performed using the statannotations (0.4.4) Python package^1^.

To evaluate the utility of the patient-level uncertainty, we performed multiple experiments described in the literature. First, similar to Hoebel et al. ^20^, we examined the linear relationship between the DSC performance and uncertainty estimates by quantifying the Pearson correlation coefficient between the certainty defined as negative uncertainty and DSC values. We developed a linear regression model with the HECKTOR DSC values as the independent variables and uncertainty values as the dependent variables. We then defined a threshold between uncertain and certain segmentations as the uncertainty value that the linear model predicted for 0.61 DSC. The threshold value was selected at 0.61 DSC since it represents the average interobserver variability for GTVp segmentation on PET/CT data as per previous literature ^14^. The patient-level uncertainty estimates were then compared to the DSC values computed on the holdout dataset, by quantifying the following possible combinations; the model is uncertain and the segmentation is inaccurate (*n*_*i,u*_), the model is uncertain and the segmentation is accurate (*n*_*a,u*_), the model is certain and the segmentation is accurate (*n*_*a,c*_), and the model is certain and the segmentation is inaccurate (*n*_*i,c*_). We then computed the following measures proposed in Mukhoti and Gal ^45^; conditional probability that the segmentation is accurate given that the model is certain *p*(*accurate*|*certain*) and Accuracy versus Uncertainty (AvU), which are defined as:

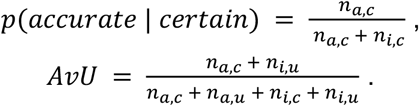

Additionally, we also report the probability that the segmentation is inaccurate given that the model is uncertain:

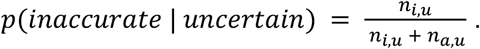

We also examined uncertainty-based referral simulation that is common in uncertainty-aware classification tasks ^39,46,47^. In the batch referral process, each patient is assigned an uncertainty score using one of the uncertainty measures and the patients are sorted based on the score. Then, the patients are removed from the set, one at a time, beginning from the highest uncertainty score, and after each removal, i.e., simulated referral, the performance measures are computed on the remaining set of patients. This process simulates a scenario where the patients for which the model has high uncertainty are referred for an expert for manual verification and/or correction, while expecting higher performance for the remaining patients. This process is repeated until 10% of the patients were remaining, as we observed that the performance measure estimates increase in stochasticity with fewer patients. As a summary score, we evaluate the area under the referral curve with Dice similarity coefficient (R-DSC AUC), in a similar manner as performed by Band et al. ^46^ with accuracy.

In addition, we evaluate the uncertainty-based referral with an instance-based process, where scans are rejected according to a predetermined uncertainty-threshold calculated from validation data. This analysis is more oriented for practice, as the cases are flagged for high uncertainty with a predetermined threshold value, instead of a value corresponding to a certain percentile computed on a batch of data, i.e., the holdout test set. We examine this process with three thresholds; uncertainty at 0.80 validation DSC, uncertainty at 0.85 validation DSC, and uncertainty at 0.90 validation DSC. As the instance referral process does not control the number of patients, but we would like for the model to confidently and accurately segment as many patients as possible, we also report the number of patients considered as certain in the instance referral process analysis.

## Results

### Segmentation performance

As for the overall segmentation performance, the MC Dropout Ensemble had a DSC value of 0.822 (SEM: 0.012) on the validation data and 0.776 on the holdout data. The Deep Ensemble DSC was found to be 0.818 (SEM: 0.014) on the validation data and 0.767 on the holdout data. As for the MSD metric, the MC Dropout Ensemble had 1.267 mm (SEM: 0.108) on the validation data and 1.703 mm on the holdout data, whereas the Deep Ensemble had 1.271 mm (SEM: 0.110) on the validation data and 1.717 mm on the holdout data. For the MC Dropout Ensemble and the Deep Ensemble, the 95HD metric was 3.658 mm (SEM: 0.423) and 3.640 mm (SEM: 0.433) on the validation data, and 5.385 mm and 5.477 mm on the holdout data, respectively. There was a statistically significant difference between the Deep Ensemble and the MC Dropout Ensemble for all the metrics on the holdout set. Overall segmentation performance for the holdout data is illustrated in **Figure 1**.

**Figure 1:**
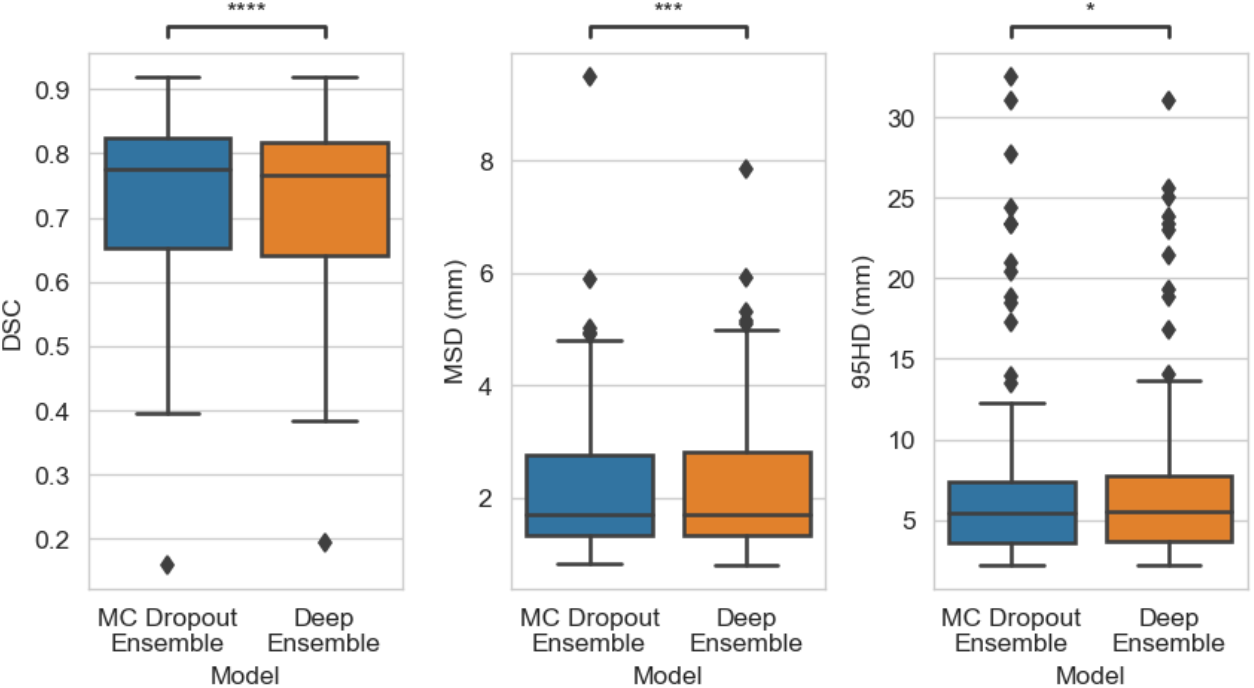
Boxplot of Dice Coefficient Score (DSC), mean surface distance (MSD), and Hausdorff distance at 95% (95HD) performance on the external dataset (N=67). Statiscal significance is measured using the Wilcoxon signed-rank test. Comparision symbols: ns(p > 0.05),*(p ≤ 0.05;),* *(p ≤ 0.01), * * *(p ≤ 1e-4), * * * *(p ≤ 1e-5).

### Uncertainty estimation

The categorized uncertain/certain cases were compared to the ground truth DSC values using the holdout set. The MC Dropout Ensemble had the highest p(uncertain | inaccurate) with *U*_*E*_, p(accurate | certain) with *U*_*CV*_, and AvU with *U*_*CV*_, with the values of 0.921, 0.800, and 0.866, respectively. The Deep Ensemble had highest p(inaccurate | uncertain), p(accurate|certain), and AvU values of 0.909, 0.750, and 0.866 with *U*_*E*_, *U*_*MI*_ and *U*_*CV*_, respectively. *U*_*E*_ had the worst AvU values of 0.582 and 0.657 for MC Dropout Ensemble and Deep Ensemble, respectively, while *U*_*MI*_ and *U*_*DR*_ had comparable AvU of 0.851 for both models. Full results are shown in **Figure 2** and **Table 1**. From the linear correlation analysis between DSC and certainty, −*U*_*CV*_ had a correlation of ρ = 0.718 (p = 7.89e-12) and ρ = 0.720 (p = 6.64e-12) for MC Dropout Ensemble and Deep Ensemble, respectively. −*U*_*E*_ had a correlation value to the DSC with ρ = -0.043 (p = 7.32e-01) and ρ = 0.465 (p = 7.24e-05), for MC Dropout Ensemble and Deep Ensemble, respectively. −*U*_*p*_ had a correlation value to the DSC with ρ = 0.704 (p = 2.99e-11) and ρ = 0.676 (p = 3.47e-10), for MC Dropout Ensemble and Deep Ensemble, respectively. −*U*_*MI*_ had a correlation value to the DSC with ρ = 0.676 (p = 3.36e-11) and ρ = 0.623 (p = 1.84e-08), for MC Dropout Ensemble and Deep Ensemble, respectively. −*U*_*DR*_ had a correlation value of ρ = 0.698 (p = 5.41e-11) and ρ = 0.704 (p = 3.13e-11) for MC Dropout Ensemble and Deep Ensemble, respectively.

**Table 1:**
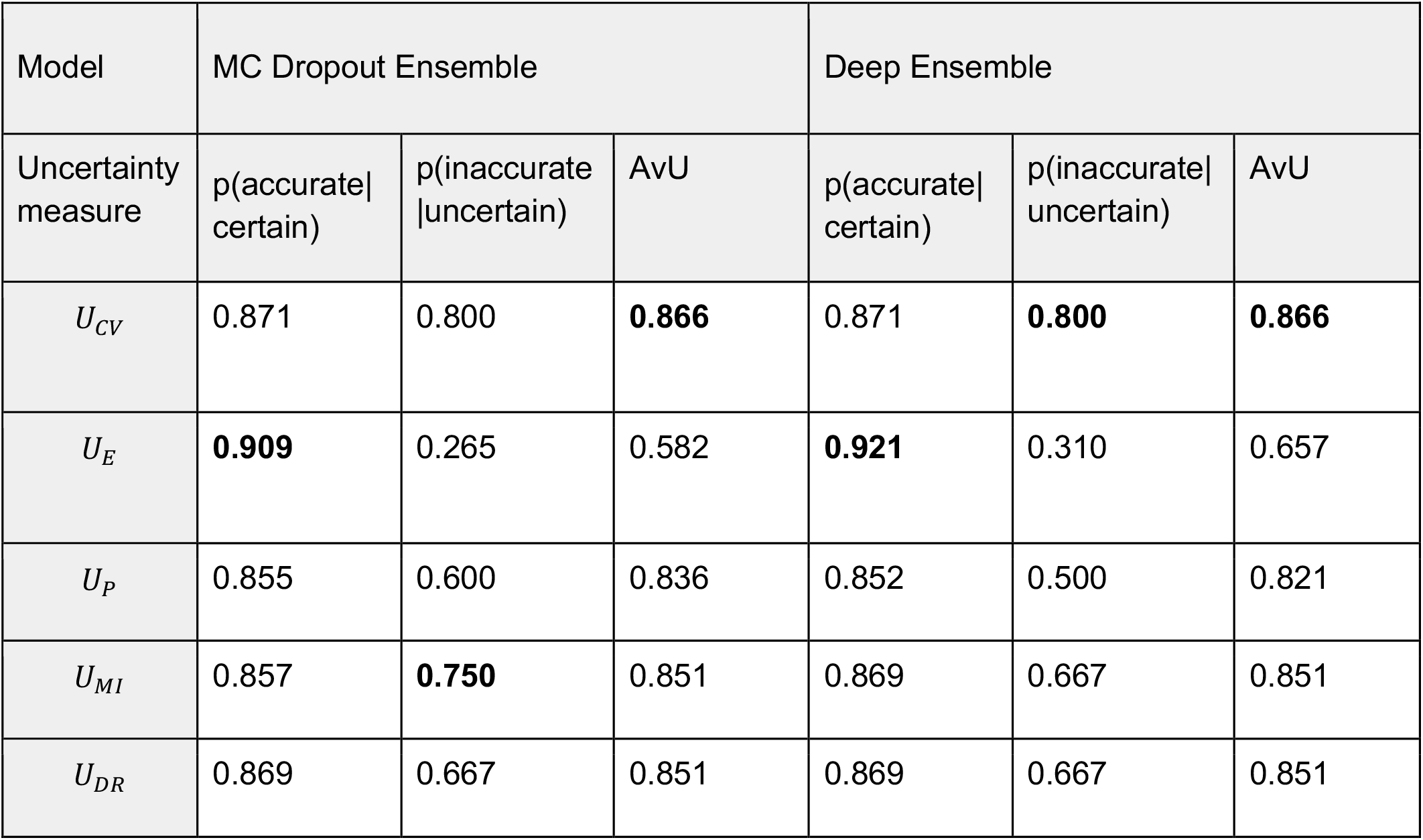
Conditional probabilities for accurate and certain cases, inaccurate and uncertain, and accuracy vs uncertainty (AvU). Accurate/inaccurate is determined by the 0.61 DSC threshold and certain/uncertain is determined by the predicted confidence threshold from 0.61 validation DSC. Best results are bolded.

**Figure 2:**
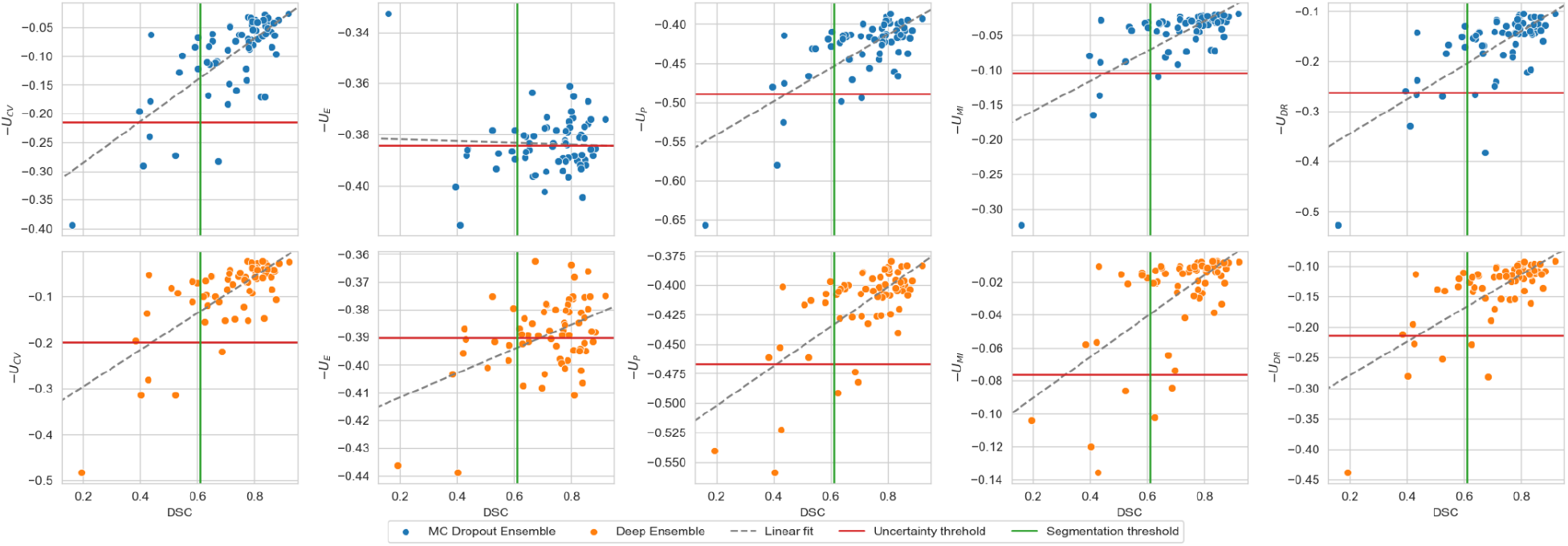
Scatterplot and linear fit of the test set Dice similarity coefficient (DSC) and model certainty based from coefficient of variation (*U*_*CV*_), expected entropy (*U*_*E*_), structure predictive entropy (*U*_*p*_), mutual information (*U*_*MI*_) and Dice-risk (*U*_*DR*_). The segmentation and uncertainty thresholds are drawn at the interobserver variability value of 0.61 DSC and at the predicted certainty value of 0.61 validation DSC, respectively.

When simulating the batch referral process on the holdout set, by rejecting the most uncertain scans up to 90% of the total number of scans, it turned out that the coefficient of variation had the highest R-DSC AUC and structure expected entropy had the lowest R-DSC AUC for both of the models. During the referral process, all the uncertainty measures generally increased the performance, except for the structure expected entropy with MC Dropout Ensemble, as the DSC decreased under the initial, i.e., full holdout set, performance around 35% and past 85% referred cases. The R-DSC AUC values for the different uncertainty measures and models are presented in **Table 2** and referral curves are shown in **Figure 3**, which also illustrates the effect of batch referral on MSD and 95HD.

**Table 2:** The area under the referral curve with Dice similarity coefficient of the models and uncertainty measures. The values are computed in the batch-based referral process and interpreted as higher is better. Best results are bolded.

| Model | $U_{CV}$ | $U_E$ | $U_P$ | $U_{MI}$ | $U_{DR}$ |
| --- | --- | --- | --- | --- | --- |
| Deep Ensemble | **0.782** | 0.752 | 0.771 | 0.772 | 0.769 |
| MC Dropout Ensemble | **0.783** | 0.734 | 0.774 | 0.778 | 0.775 |

**Figure 3:**
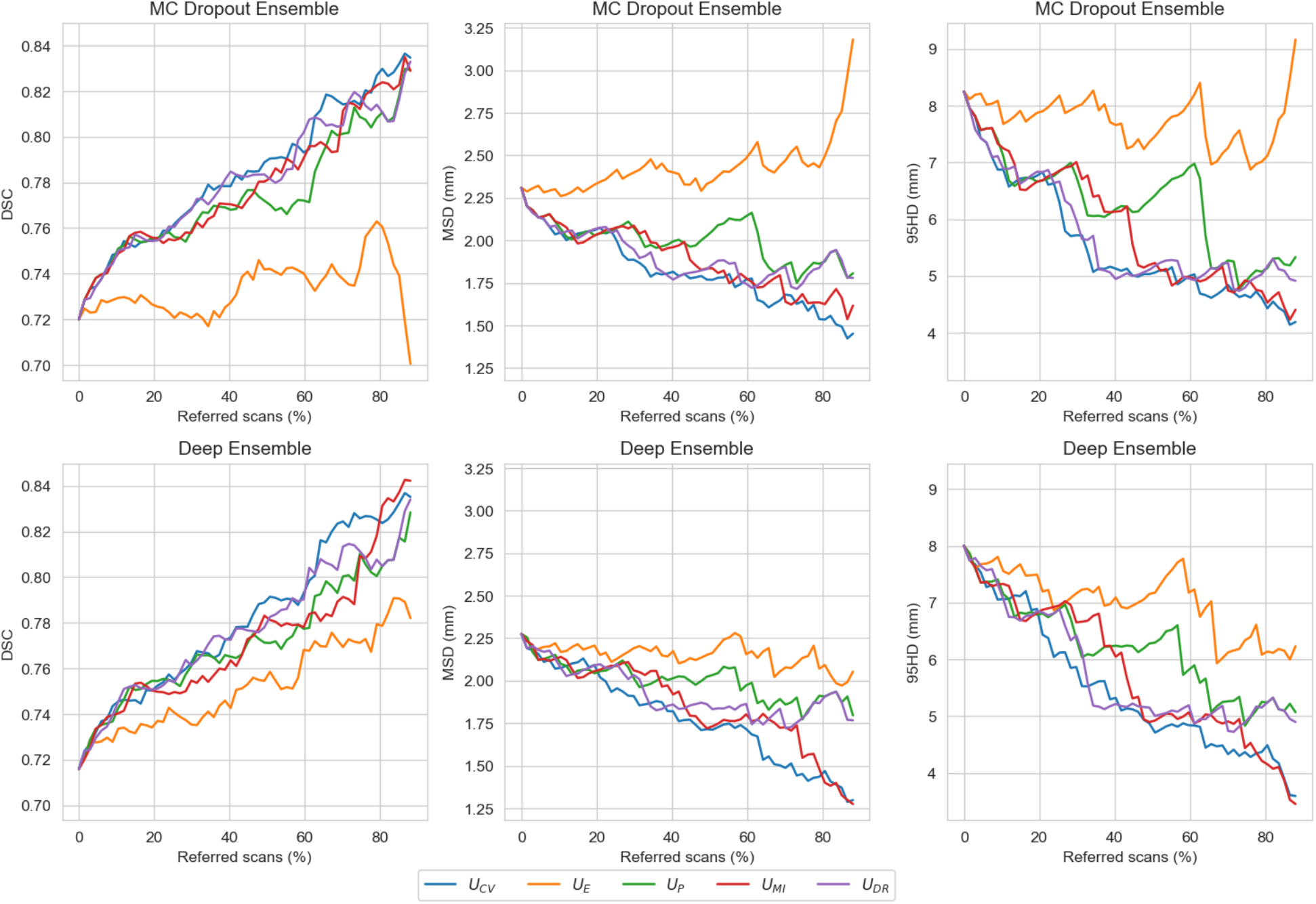
Model performance measured with Dice similarity coefficient (DSC), mean surface distance (MSD), and Hausdorff distance at 95% (95HD), on various referral levels in the batch referral process, when referring the most uncertain scans, based on coefficient of variation, structure expected entropy, structure predictive entropy, structure mutual information, and Dice-risk, up to 90% of the total scans.

For the instance-based referral process measured with a validation DSC uncertainty threshold of 0.80 all of the uncertainty measures kept all patients and full dataset performance for the Deep Ensemble while for MC Dropout Ensemble the *U*_*p*_ and *U*_*MI*_ removed a single patient and improved DSC to 0.728. For the 0.85 validation DSC threshold, the MC Dropout Ensemble had the highest DSC value of 0.769 with the *U*_*CV*_ and 47 patients retained and for the Deep Ensemble the *U*_*DR*_ had the highest DSC value of 0.753 with 52 patients retained. For the 0.9 validation DSC threshold, the MC Dropout Ensemble had the highest DSC value of 0.876 with the *U*_*CV*_ *and* 2 patients retained and for the Deep Ensemble the *U*_*DR*_ had the highest DSC value of 0.808 with 11 patients retained. For both of the models and with the 0.85 and 0.90 expected DSC uncertainty thresholds, *U*_*p*_ retained the most patients with 55, and 12 for the MC Dropout Ensemble, and with 56, and 13 for the Deep Ensemble. Full results for all uncertainty measures for both models and all thresholds are shown in **Table 3**.

**Table 3:** DSC performance and number of patients retained after referring to uncertain cases based on validation DSC. Best results for each threshold and model are bolded.

|  | Model | MC Dropout Ensemble |  | Deep Ensemble |  |
| --- | --- | --- | --- | --- | --- |
| Validation DSC | Uncertainty measure | DSC | Number of patients retained | DSC | Number of patients retained |
| 0.80 | $U_{CV}$ | 0.720 | **67** | **0.716** | **67** |
| | $U_E$ | 0.720 | **67** | **0.716** | **67** |
| | $U_P$ | **0.728** | 66 | **0.716** | **67** |
| | $U_{MI}$ | **0.728** | 66 | **0.716** | **67** |
| | $U_{DR}$ | 0.720 | **67** | **0.716** | **67** |
| 0.85 | $U_{CV}$ | **0.769** | 47 | 0.752 | 53 |
| | $U_E$ | - | 0 | - | 0 |
| | $U_P$ | 0.754 | **55** | 0.751 | **56** |
| | $U_{MI}$ | 0.755 | 51 | 0.750 | 48 |
| | $U_{DR}$ | 0.755 | 55 | **0.753** | 52 |
| 0.90 | $U_{CV}$ | **0.876** | 2 | - | 0 |
| | $U_E$ | - | 0 | - | 0 |
| | $U_P$ | 0.807 | **12** | 0.805 | **13** |
| | $U_{MI}$ | 0.835 | 9 | - | 0 |
| | $U_{DR}$ | 0.807 | 11 | **0.808** | 11 |

When visually examining the predictive entropy, mutual information, and expected entropy for both MC Dropout Ensemble and Deep Ensemble models, the uncertainty is highest around the edges of the predicted segmentation mask for all the measures. Mutual information is mainly focused on the edges with the inner volume having high confidence, while expected entropy demonstrates moderate uncertainty near the inner volume. Full visual comparison of axial slices is shown in **Figure 4**. Additional in-depth qualitative analysis of uncertainty maps for select cases are shown in **Appendix B**.

**Figure 4:**
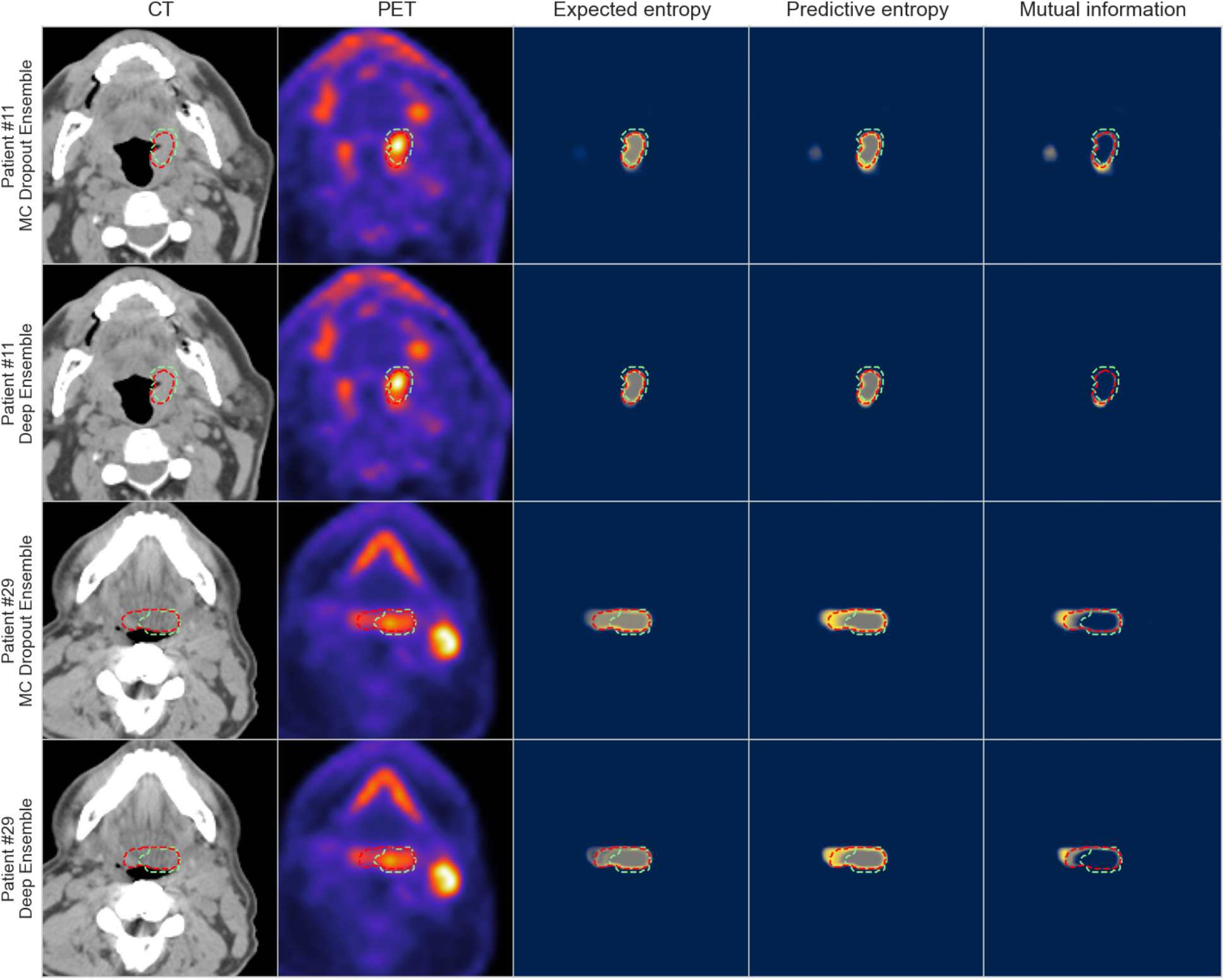
Visualization of model segmentation and uncertainty maps for two patients. The columns illustrate in order: computed tomography (CT), positron emission tomography (PET), expected entropy, predictive entropy and mutual information. The model and expert segmentations are superimposed in red and green, respectively. Rows one and three contain the results for MC Dropout Ensemble while rows two and four contain the results for the Deep Ensemble. Blue, gray, and yellow colors in uncertainty maps correspond to low, medium, and high model uncertainty, respectively.

## Discussion

In this study, we have systematically investigated two established variations of Bayesian inference in DL for OPC GTVp segmentation, namely the MC Dropout Ensemble and the Deep Ensemble. Through experimentation with a multitude of uncertainty quantification measures (coefficient of variation, structure expected entropy, structure predictive entropy, structure mutual information, and Dice-risk) we compare and contrast differences between these approaches. Given the relative sparsity of existing literature for uncertainty estimation in OPC-related segmentation, our results act as an essential first benchmarking step towards a deeper understanding and further implementation of these techniques for clinically applicable radiotherapy segmentation workflows.

Both of the evaluated probabilistic DL methods had similar segmentation performance with larger DSC scores compared to expected average expert interobserver variation. While statistical testing did demonstrate some differences between the methods, it should be noted that these minor differences are likely not clinically meaningful. Generally speaking, the state-of-the-art performance for the DL-based PET/CT OPC GTVp segmentation has remained mostly stagnant over the past few years, with external validation results being within a similar range as ours ^16,37^. This is likely secondary to the already established large interobserver variability in OPC tumor-related segmentation. This further emphasizes the need for methods to provide clinicians with uncertainty estimates that could be used to further guide their clinical decision making.

In terms of the utility of uncertainty for estimating the patient-level performance, both models had the highest Accuracy versus Uncertainty metric, i.e., the proportion of correctly identified high and low DSC cases, with the coefficient of variation uncertainty measure. Moreover, the AvU value was the same with the models when using coefficient of variation, structural mutual information, and Dice-risk, which suggests that the Deep Ensemble can be considered to be as accurate in the uncertainty estimation as the MC Dropout Ensemble, while requiring less computational resources. Indeed, our results are well aligned with a recent large scale study of Bayesian DL, which showed that the Deep Ensemble is a competitive approximation for Bayesian inference ^35^. As our uncertainty quantification methods require no modifications to the training of these models, and as the Deep Ensemble has been a popular approach for PET/CT OPC tumor segmentation (17 out of the 22 teams participating in the 2021 HECKTOR segmentation challenge utilized model ensembling ^16^), it is straightforward to apply our uncertainty quantification approaches for existing models to enable the model confidence to be used in practice.

Among all the uncertainty measures investigated, the coefficient of variation had generally favorable performance in terms of the accuracy of uncertainty-based segmentation performance prediction and both referral processes. Interestingly, the structure expected entropy, used in several previous works, turned out to be the worst uncertainty measure in all the experiments. This finding suggests that most of the uncertainty in this task is related to the model uncertainty, as the expected entropy has been described to capture the aleatoric component of uncertainty ^46^. In the instance referral process, designed to simulate the real use-case conditions, the Deep Ensemble benefited most from the use of our Dice-risk uncertainty measure, whereas the MC Dropout Ensemble benefited mostly from the coefficient of variation. However, the coefficient of variation seems to produce conservative estimates as it also referred more patients when compared to the other uncertainty measures. Finally, it is worth noting our DSC-based Dice-risk uncertainty measure had comparable performance to these measures, but was outperformed by the coefficient of variation in many experiments. The expected conditional risk used to develop the measure could be extended to reflect clinical preference or risk-averseness, such as the shape, size, and location of the model output segmentation, to fine-tune the uncertainty estimation for the specific OPC tumor segmentation task.

From our qualitative analysis, both of the methods produced uncertain voxels mainly on the edges of the predicted segmentation mask for the expected entropy, predictive entropy, and mutual information including both the aleatoric and epistemic components of the uncertainty. This is likely reinforced by the selection of Dice loss for the objective. When comparing the models, the MC Dropout Ensemble method provided a smoother uncertainty gradient and more variation in the uncertainty, likely due to providing 300 samples per voxel compared to the five samples of Deep Ensemble. Moreover, a key takeaway from additional qualitative analysis includes a general overemphasis of PET signal by the models that could lead to erroneous predictions and uncertainty quantification. This however, is not necessarily unexpected, since studies of PET/CT auto-segmentation have demonstrated that models generally utilize PET signal to a higher degree in model predictions compared to CT signal ^50,51^.

Lei et al. ^31^, Tang et al. ^30^, and van Rooij et al. ^32^ are among the few studies that have investigated segmentation related uncertainties in HNSCC. Similar to our work, these studies utilized ensembling (Lei et al.) and MC Dropout (Tang et al., van Rooij et al.) to segment nasopharyngeal cancer tumors and organs at risk on CT images, respectively. Moreover, in the only currently published study on the topic of uncertainty estimation in OPC GTVp segmentation, De Biase et al. ^33^ proposed a novel DL-based method using PET/CT images that generated probability maps for capturing the model uncertainty. The sequences of three consecutive 2-dimensional slices and the corresponding tumor segmentations were used as inputs to a model that leveraged inter/intra-slice context using attention mechanisms and recurrent neural network architectures. In their study, ensembling was used to derive probability maps rather than uncertainty maps, whereupon the authors experimented with different probability thresholds corresponding to areas of higher or lower agreement among the trained models. Our study acts as an important adjunct to De Bias et al., as the various methodologies investigated herein could be coupled with their proposed clinical solution.

There are some limitations in our study. Firstly, we examined only two commonly used probabilistic DL models and focused on five uncertainty measures. These methods and four of the uncertainty measures were selected due to their relative prevalence in existing literature and were thus deemed as an important starting point for exploring uncertainty estimation in OPC tumor-related segmentation. Secondly, we have utilized a relatively limited sample size for model training and evaluation. However, this study contains a robust training set from multiple institutions as supplied by the de-facto standard data science competition for OPC segmentation (i.e., HECKTOR) with external validation through our own institutional holdout dataset. Moreover, we have chosen to utilize bounding boxes around the GTVp, as was performed for 2021 HECKTOR Challenge, in order to simplify the segmentation problem and focus on the exploration of uncertainty estimation; future studies should attempt the integration of uncertainty estimation into fully developed OPC segmentation workflows that can be applied to “as encountered” PET/CT images. Thirdly, we have limited our investigation to the primary tumor, and not investigated nodal metastasis in this study, but as the newer editions of the HECKTOR Challenge includes these regions of interest, the incorporation of nodal metastasis should be the focus of future studies. Fourthly, we have only investigated segmentations generated by a single observer for each scan, but the influence of multi-observer segmentations on uncertainty estimates is a future research direction. Finally, we have chosen to squarely focus on PET/CT as an imaging modality due to its ubiquity in OPC GTVp segmentation workflows. However, it is known that different imaging modalities (e.g., magnetic resonance imaging, contrast enhanced CT) can provide complementary information for OPC tumor segmentation ^52^, and the combination of multiple image inputs may affect auto-segmentation model outputs ^10,50,53^. Therefore, future research should investigate how results differ for models using alternative imaging modalities and the impacts of individual channel inputs on the uncertainty estimation.

## Conclusion

We applied probabilistic DL models for OPC GTVp segmentation using multimodal large-scale datasets in order to evaluate the utility of uncertainty estimation through a variety of uncertainty measures. We found that regardless of the uncertainty measure applied both of the probabilistic DL methods (Deep Ensemble and MC Dropout Ensemble) provided similar utility in terms of predicting segmentation quality and referral performance; due to its slightly lower computational cost and greater ubiquity, Deep Ensemble may be preferable to MC Dropout Ensemble. Notably, coefficient of variation had overall favorable performance for both models so may be ideal as an uncertainty measure. While research in uncertainty estimation for OPC GTVp auto-segmentation is in its nascent stage, we anticipate that uncertainty estimation will become increasingly important as these AI-based technologies begin to enter clinical workflows. Therefore our benchmarking study is a crucial first-step towards a wider adoption and exploration of these techniques. Future studies should investigate further uncertainty quantification methodology, larger sample sizes, additional relevant segmentation targets (i.e., metastatic lymph nodes), and incorporation of additional imaging modalities.

## Data Availability

All data produced in the present study are available upon reasonable request to the authors.

## Funding Acknowledgements

The work of Joel Jaskari, Jaakko Sahlsten, and Kimmo K. Kaski was supported in part by the Academy of Finland under Project 345449. Kareem A. Wahid is supported by the Dr. John J. Kopchick Fellowship through The University of Texas MD Anderson UTHealth Graduate School of Biomedical Sciences, the American Legion Auxiliary Fellowship in Cancer Research, and an NIH/National Institute for Dental and Craniofacial Research (NIDCR) F31 fellowship (1 F31DE031502-01). Benjamin H. Kann is supported by an NIH/National Institute for Dental and Craniofacial Research (NIDCR) K08 Grant (K08DE030216). Clifton D. Fuller receives related grant support from the NCI NRSA Image Guided Cancer Therapy Training Program (T32CA261856), as well as additional unrelated salary/effort support from NIH institutes. Dr. Fuller receives grant and infrastructure support from MD Anderson Cancer Center via: the Charles and Daneen Stiefel Center for Head and Neck Cancer Oropharyngeal Cancer Research Program; the Program in Image-guided Cancer Therapy; and the NIH/NCI Cancer Center Support Grant (CCSG) Radiation Oncology and Cancer Imaging Program (P30CA016672). Dr. Fuller has received unrelated direct industry grant/in-kind support, honoraria, and travel funding from Elekta AB.

## Appendix A: Supplementary Methods

Additional details on the MDA external validation dataset are provided here. For the 67 patients included, FDG-PET/CT scans were acquired from various GE Medical Systems scanners. Specifically, Discovery RX (n=27), Discovery STE (n=26), Discovery ST (n=12) and Discovery HR (n=2) models were used. Image acquisition parameters are shown in **Table A1**. A 90-minute uptake period of rest was used for all patients. Attenuation corrected images were reconstructed using an ordered subset expectation maximization (OSEM) iterative algorithm (2 iterations, 18-24 subsets, 5mm Gaussian filter).

**Table A1:** PET/CT image acquisition parameters. All values were the same across all patients unless a parenthesis is shown, where the median and range are displayed. * only apply to CT data; ** only apply to PET data.

| Acquisition Parameter | CT | PET |
| --- | --- | --- |
| In-plane resolution (mm) | 0.98 | 5.47 |
| Slice thickness (mm) | 3.75 | 3.27 |
| Exposure time (ms)* | 566 (500-676) | NA |
| X-ray tube current (mA)* | 200 (100-296) | NA |
| KVP (kV)* | 100 (100-120) | NA |
| Dose (Mbq)** | NA | 375 (281-729) |

## Appendix B: Additional Qualitative Analysis

In this section, we present qualitative results for select cases in the MDA holdout dataset. Specifically, we describe our interpretations of model predictions and uncertainty maps relative to ground truth across multiple axial image slices, similar to how a case would be reviewed in the clinic. For simplicity, we only describe results of the MD Dropout Ensemble model. “High” and “low” values are relative to median values described in the main text (e.g., high DSC is greater than 0.61).

### Case 1: Low performance, high certainty

Here we describe a case with low DSC (0.43) but high certainty (−*U*_*p*_ =−0.43). Axial slice representations from superior to inferior slices for this case are shown in **Figure B1**. As can be seen in the inferior slices (slice 54), there is initially a relatively large degree of uncertainty about the beginning of the prediction. Subsequently (slice 66), the model correctly predicts the tumor at the left base of tongue, with a simultaneous region of uncertainty appearing at the right base of tongue, likely secondary to the high PET signal causing a potential area of false positivity. This false positive PET signal is not ultimately included in the predicted segmentation mask, which in this case is seen as a desired outcome. More superiorly (slice 79), in terms of uncertainty and the resultant prediction, the model seems to have erroneously localized to the hyper-metabolic core of the primary tumor. Finally, at the most superior slices (slice 90), it is noted that there was metal streak artifact induced by dental hardware, which may have interfered with model inference and subsequent uncertainty quantification, as no prediction was generated. Main takeaways from this case include the model overemphasizing PET signal (which has been previously noted in PET/CT auto-segmentation models) which is also reflected in the resultant uncertainty measures. Moreover, the image artifact may also impact performance and uncertainty estimation.

**Figure B1:**
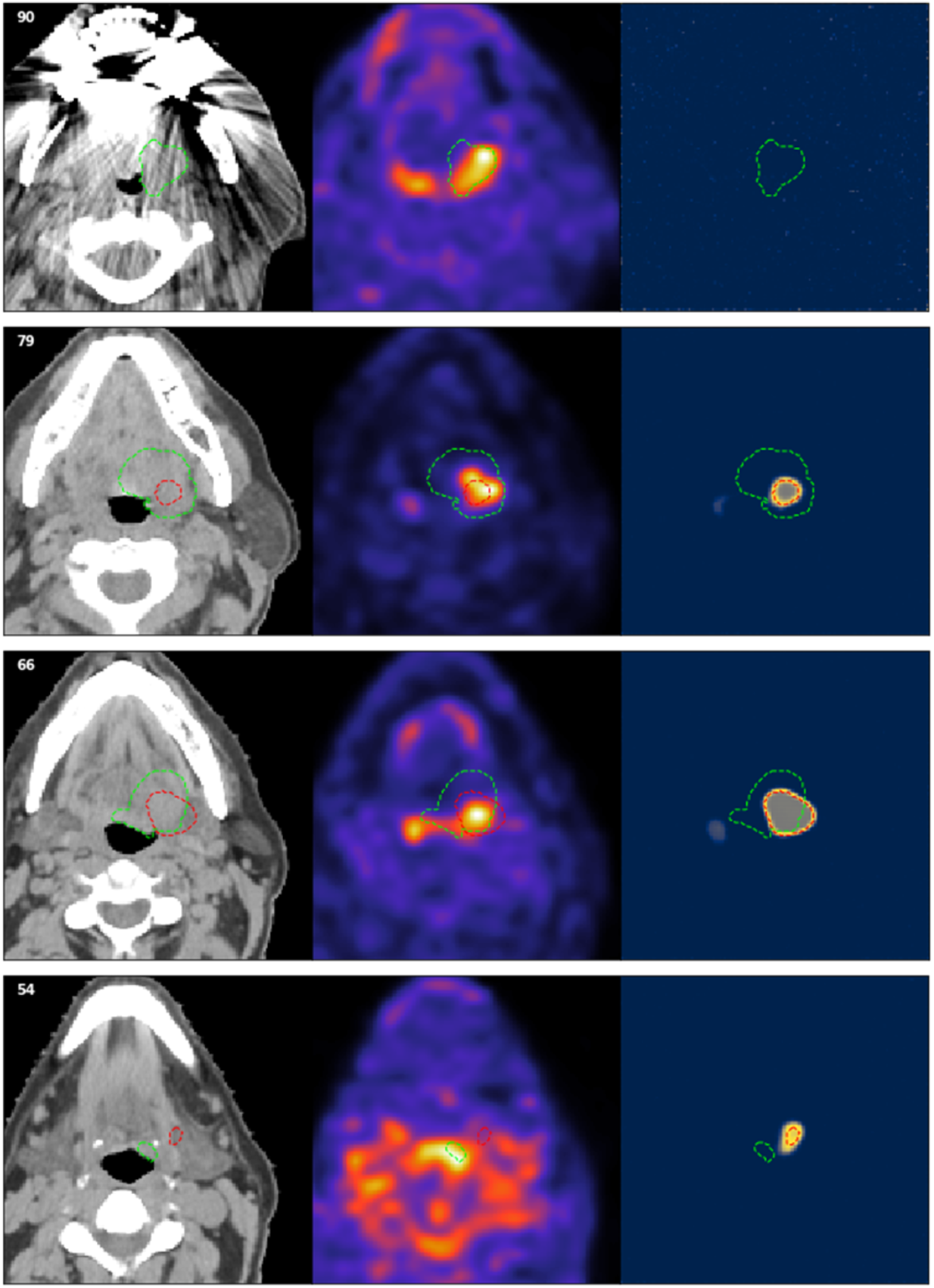
Additional qualitative investigation of a case with low performance and high certainty. Number in top left corner = slice number; green dotted outline = ground-truth segmentation, red dotted outline = predicted segmentation. Blue, gray, and yellow colors in uncertainty maps correspond to low, medium, and high model uncertainty, respectively.

### Case 2: High performance, low certainty

Here we describe a case with high DSC (0.64) but low certainty (−*U*_*p*_ =−0.5). Axial slice representations from superior to inferior slices for this case are shown in **Figure B2**. In the inferior-most slices (slices 18-27), uncertainty is noted near the larynx, likely a byproduct of high PET signal. As before, this false positive PET signal is not ultimately included in the predicted segmentation mask, which in this case is seen as a desired outcome. More superiorly (slice 61), the model begins to predict a segmentation on only the right side of the base of tongue, when in reality the ground-truth is a bilateral segmentation. Importantly, the model starts to note uncertainty on the contralateral part of the image, which is a desired outcome. As we move further superiorly towards the tonsils (slice 70) the tumor begins to exhibit an uncommon presentation (discontinuous fragment, bilateral in both tonsils), but the prediction better starts to approximate the ground-truth; the uncertainty previously demonstrated at the contralateral side (left) is still present but has now started to become included in the predicted segmentation. Continuing superiorly (slice 78), there is still high uncertainty in the discontinuous fragment but the model is able to generate a reasonable prediction, however the model eventually starts to generate an implausible prediction in an air space (slice 85) as the prediction begins to generate a bilateral segmentation erroneously. As with the previous case, towards the superior-most part of the image (slices 90-94), metal streak artifact induced by dental hardware may alter the predictions and uncertainty estimation; notably, the prediction ignores the false positive PET signal. Main takeaways from this case include uncommon tumor presentations (e.g., fragmentation of tumor from one continuous piece to two pieces) may present issues in generating prediction and uncertainty. Moreover, as before, image artifacts may impact predictions and uncertainty estimation.

**Figure B2:**
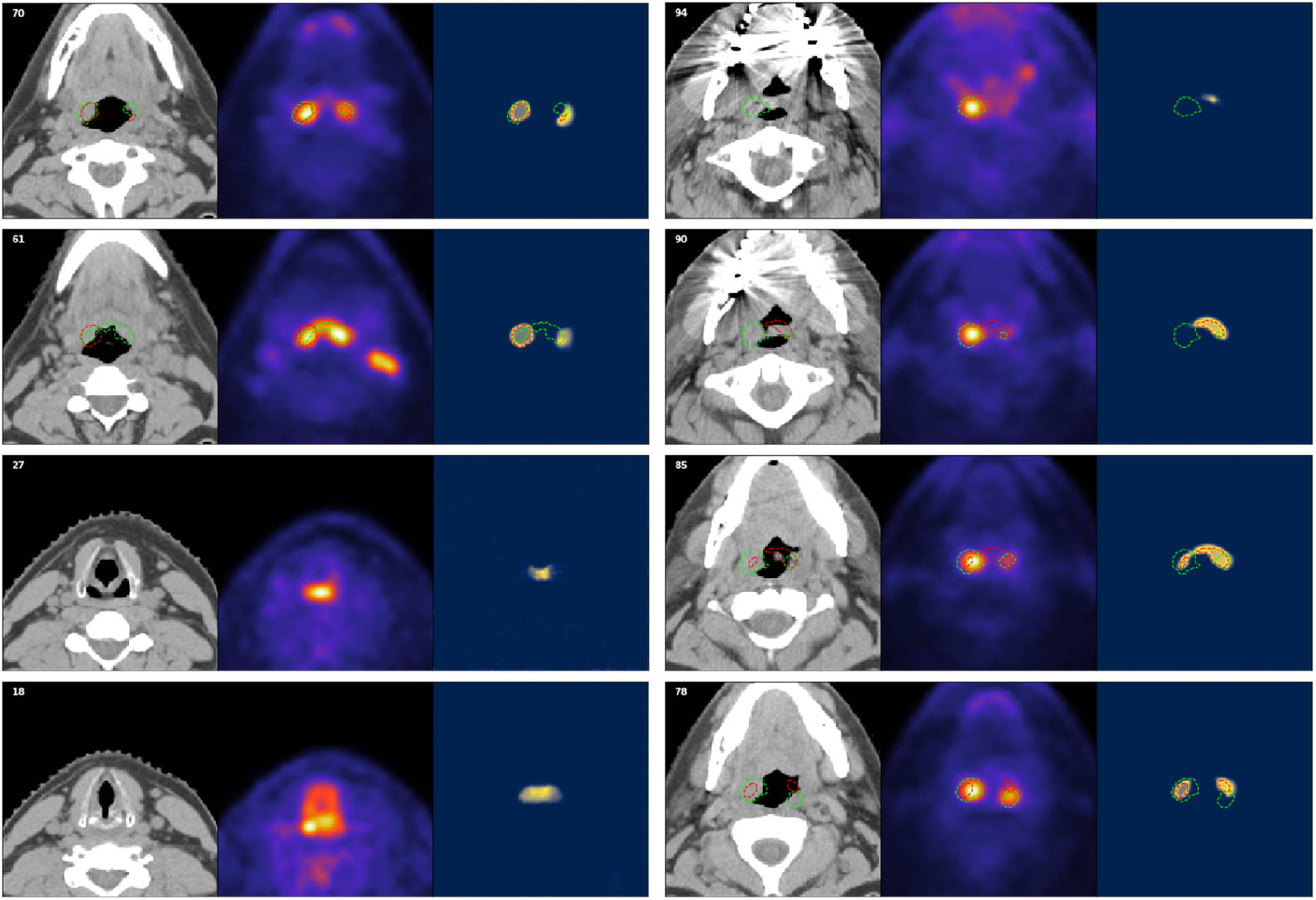
Additional qualitative investigation of a case with high performance and low certainty. Number in top left corner = slice number; green dotted outline = ground-truth segmentation, red dotted outline = predicted segmentation. Blue, gray, and yellow colors in uncertainty maps correspond to low, medium, and high model uncertainty, respectively.

### Case 3: Contralateral uncertainty

Here we describe an interesting case with high DSC (0.64) and high certainty (−*U*_*p*_ = −0.41). Axial slice representations from superior to inferior slices for this case are shown in **Figure B3**. At the inferior slice (slice 60), the model generates the prediction correctly at the left tonsil but starts to note uncertainty at the contralateral tonsil. Subsequently, at the more superior slice (slice 70) the contralateral portion is revealed as part of the ground truth segmentation. The model is still uncertain about the area and ultimately does not include it as part of the prediction. In other words, the contralateral uncertainty indicates a false negative area that the model is uncertain about. In a clinical workflow this would correspond to an area the clinician could choose to further investigate.

**Figure B3:**
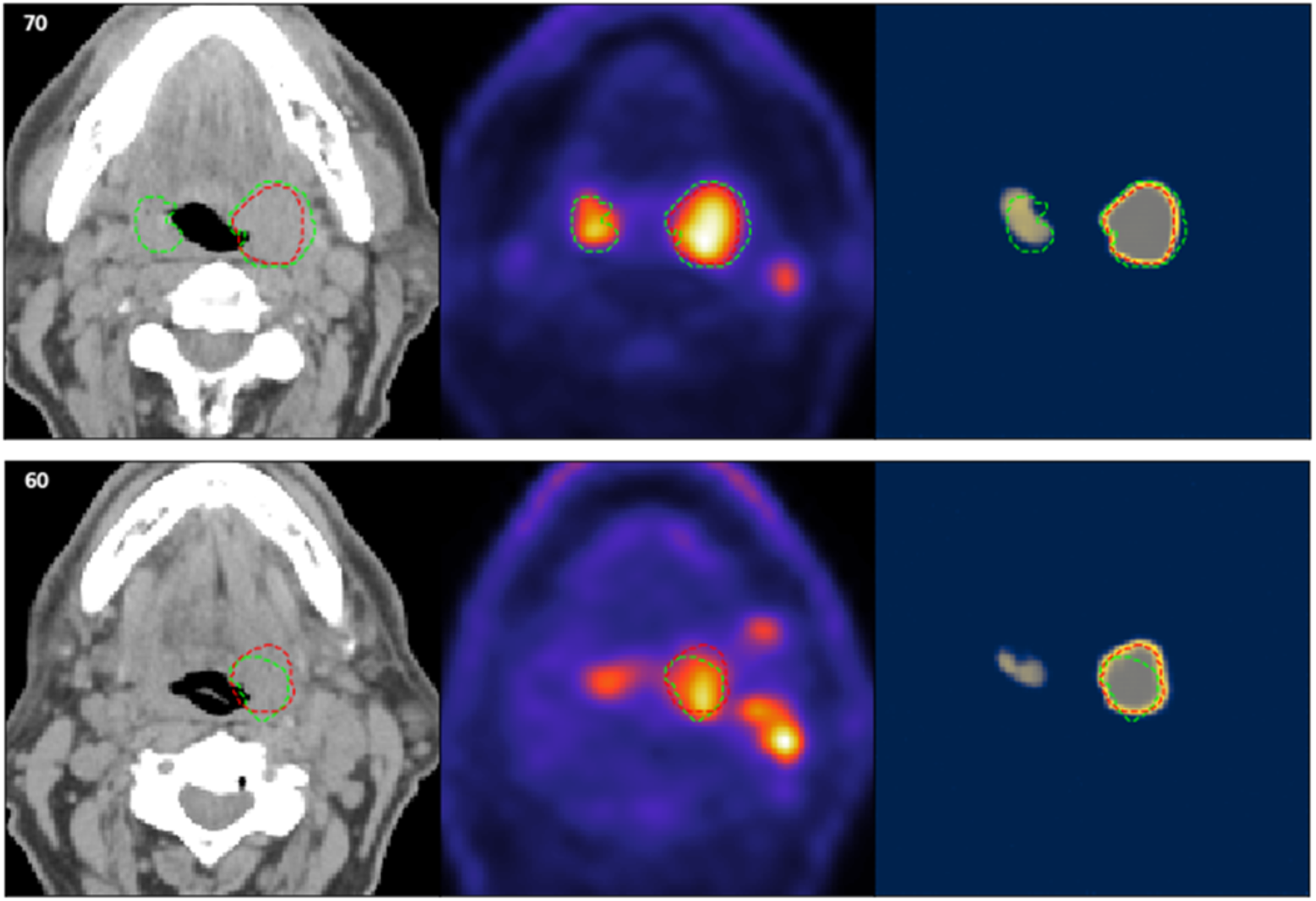
Additional qualitative investigation of a case with contralateral uncertainty. Number in top left corner = slice number; green dotted outline = ground-truth segmentation, red dotted outline = predicted segmentation. Blue, gray, and yellow colors in uncertainty maps correspond to low, medium, and high model uncertainty, respectively.

### Case 4: Nodal uncertainty

Here we describe an interesting case with high DSC (0.71) and high certainty (−*U*_*p*_ = −0.44). Axial slice representations from superior to inferior slices for this case are shown in **Figure B4**. In the inferior-most slice (slice 22) there is noted uncertainty in the area of high PET signal (likely spurious signal), which is not included in the prediction, which in this case is seen as a desired outcome. More superiorly (slices 61-70) a metastatic lymph node is present on the right side of the image; there is corresponding noted uncertainty about this area and it is ultimately not included in the prediction. The model is able to generate a prediction for the right base of tongue tumor without issues. Notably, as observed through the majority of other cases, metastatic lymph nodes are normally not considered by the model at all, likely due to the often large geometric distances between the nodal metastases and the primary tumors. In this case the node exhibits features (i.e. high PET signal) in close proximity to the primary tumor, which could have led to the model uncertainty about this prediction.

**Figure B4:**
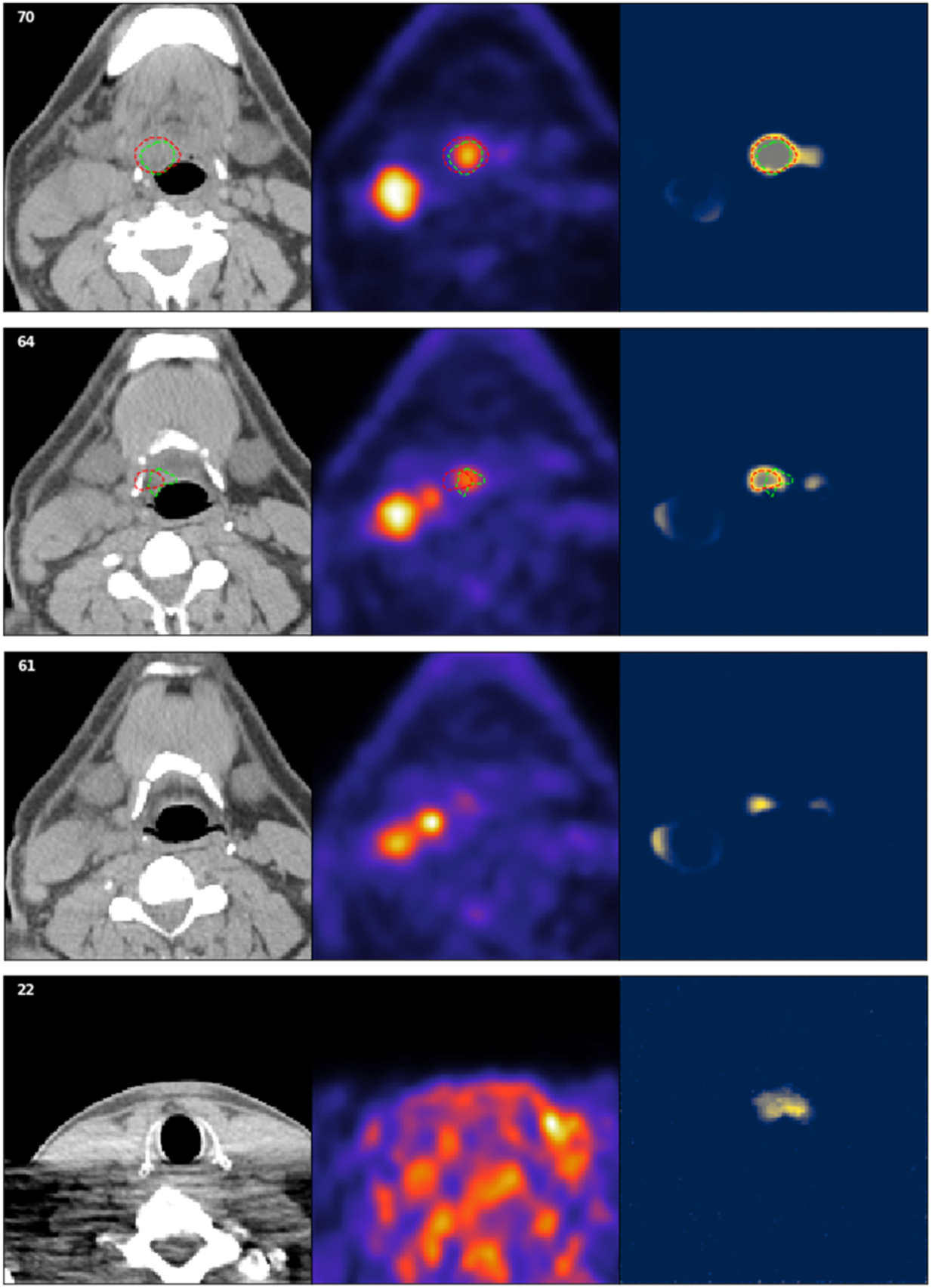
Additional qualitative investigation of a case with nodal uncertainty. Number in top left corner = slice number; green dotted outline = ground-truth segmentation, red dotted outline = predicted segmentation. Blue, gray, and yellow colors in uncertainty maps correspond to low, medium, and high model uncertainty, respectively.

## Footnotes

1 https://github.com/trevismd/statannotations

## References

1. Bray, F., Ferlay, J., Soerjomataram, I., Siegel, R. L., Torre, L. A. & Jemal, A. Global cancer statistics 2018: GLOBOCAN estimates of incidence and mortality worldwide for 36 cancers in 185 countries. CA Cancer J. Clin. 68, 394–424 (2018).

2. Rasch, C., Steenbakkers, R. & van Herk, M. Target definition in prostate, head, and neck. Semin. Radiat. Oncol. 15, 136–145 (2005).

3. Cardenas, C. E., Blinde, S. E., Mohamed, A. S. R., Ng, S. P., Raaijmakers, C., Philippens, M., Kotte, A., Al-Mamgani, A. A., Karam, I., Thomson, D. J., Robbins, J., Newbold, K., Fuller, C. D. & Terhaard, C. Comprehensive Quantitative Evaluation of Variability in Magnetic Resonance-Guided Delineation of Oropharyngeal Gross Tumor Volumes and High-Risk Clinical Target Volumes: An R-IDEAL Stage 0 Prospective Study. Int. J. Radiat. Oncol. Biol. Phys. 113, 426–436 (2022).

4. Lin, D., Wahid, K. A., Nelms, B. E., He, R., Naser, M. A., Duke, S., Sherer, M. V., Christodouleas, J. P., Mohamed, A. S. R., Cislo, M., Murphy, J. D., Fuller, C. D. & Gillespie, E. F. E pluribus unum: prospective acceptability benchmarking from the Contouring Collaborative for Consensus in Radiation Oncology crowdsourced initiative for multiobserver segmentation. J Med Imaging (Bellingham) 10, S11903 (2023).

5. Njeh, C. F. Tumor delineation: The weakest link in the search for accuracy in radiotherapy. J. Med. Phys. 33, 136–140 (2008).

6. Segedin, B. & Petric, P. Uncertainties in target volume delineation in radiotherapy – are they relevant and what can we do about them? Radiology and Oncology 50, 254–262 Preprint at https://doi.org/10.1515/raon-2016-0023 (2016)

7. Nikolov, S., Blackwell, S., Zverovitch, A., Mendes, R., Livne, M., De Fauw, J., Patel, Y., Meyer, C., Askham, H., Romera-Paredes, B., Kelly, C., Karthikesalingam, A., Chu, C., Carnell, D., Boon, C., D’Souza, D., Moinuddin, S. A., Garie, B., McQuinlan, Y., Ireland, S., Hampton, K., Fuller, K., Montgomery, H., Rees, G., Suleyman, M., Back, T., Hughes, C. O., Ledsam, J. R. & Ronneberger, O. Clinically Applicable Segmentation of Head and Neck Anatomy for Radiotherapy: Deep Learning Algorithm Development and Validation Study. J. Med. Internet Res. 23, e26151 (2021).

8. McDonald, B. A., Cardenas, C., O’Connell, N., Ahmed, S., Naser, M. A., Wahid, K. A., Xu, J., Thill, D., Zuhour, R., Mesko, S., Augustyn, A., Buszek, S. M., Grant, S., Chapman, B. V., Bagley, A., He, R., Mohamed, A., Christodouleas, J. P., Brock, K. K. & Fuller, C. D. Investigation of autosegmentation techniques on T2-weighted MRI for off-line dose reconstruction in MR-linac Adapt to Position workflow for head and neck cancers. bioRxiv (2021). doi:10.1101/2021.09.30.21264327

9. Taku, N., Wahid, K. A., van Dijk, L. V., Sahlsten, J., Jaskari, J., Kaski, K., Fuller, C. D. & Naser, M. A. Auto-detection and segmentation of involved lymph nodes in HPV-associated oropharyngeal cancer using a convolutional deep learning neural network. Clinical and Translational Radiation Oncology 36, 47–55 (2022).

10. Wahid, K. A., Ahmed, S., He, R., van Dijk, L. V., Teuwen, J., McDonald, B. A., Salama, V., Mohamed, A. S. R., Salzillo, T., Dede, C., Taku, N., Lai, S. Y., Fuller, C. D. & Naser, M. A. Evaluation of deep learning-based multiparametric MRI oropharyngeal primary tumor auto-segmentation and investigation of input channel effects: Results from a prospective imaging registry. Clin Transl Radiat Oncol 32, 6–14 (2022).

11. Naser, M. A., van Dijk, L. V., He, R., Wahid, K. A. & Fuller, C. D. Tumor Segmentation in Patients with Head and Neck Cancers Using Deep Learning Based-on Multi-modality PET/CT Images. in Head and Neck Tumor Segmentation 85–98 (Springer International Publishing, 2021).

12. Naser, M. A., Wahid, K. A., van Dijk, L. V., He, R., Abdelaal, M. A., Dede, C., Mohamed, S. R. & Fuller, C. D. Head and Neck Cancer Primary Tumor Auto Segmentation Using Model Ensembling of Deep Learning in PET/CT Images. in Head and Neck Tumor Segmentation and Outcome Prediction 121–133 (Springer International Publishing, 2022).

13. Savjani, R. R., Lauria, M., Bose, S., Deng, J., Yuan, Y. & Andrearczyk, V. Automated Tumor Segmentation in Radiotherapy. Semin. Radiat. Oncol. 32, 319–329 (2022).

14. Oreiller, V., Andrearczyk, V., Jreige, M., Boughdad, S., Elhalawani, H., Castelli, J., Vallières, M., Zhu, S., Xie, J., Peng, Y., Iantsen, A., Hatt, M., Yuan, Y., Ma, J., Yang, X., Rao, C., Pai, S., Ghimire, K., Feng, X., Naser, M. A., Fuller, C. D., Yousefirizi, F., Rahmim, A., Chen, H., Wang, L., Prior, J. O. & Depeursinge, A. Head and neck tumor segmentation in PET/CT: The HECKTOR challenge. Med. Image Anal. 77, 102336 (2022).

15. Wahid, K. A., Glerean, E., Sahlsten, J., Jaskari, J., Kaski, K., Naser, M. A., He, R., Mohamed, A. S. R. & Fuller, C. D. Artificial Intelligence for Radiation Oncology Applications Using Public Datasets. Semin. Radiat. Oncol. 32, 400–414 (2022).

16. Andrearczyk, V., Oreiller, V., Boughdad, S., Rest, C. C. L., Elhalawani, H., Jreige, M., Prior, J. O., Vallières, M., Visvikis, D., Hatt, M. & Depeursinge, A. Overview of the HECKTOR Challenge at MICCAI 2021: Automatic Head and Neck Tumor Segmentation and Outcome Prediction in PET/CT Images. in Head and Neck Tumor Segmentation and Outcome Prediction 1–37 (Springer International Publishing, 2022).

17. Kompa, B., Snoek, J. & Beam, A. L. Second opinion needed: communicating uncertainty in medical machine learning. NPJ Digit Med 4, 4 (2021).

18. van den Berg, C. A. T. & Meliadò, E. F. Uncertainty Assessment for Deep Learning Radiotherapy Applications. Semin. Radiat. Oncol. 32, 304–318 (2022).

19. Hu, S., Worrall, D., Knegt, S., Veeling, B., Huisman, H. & Welling, M. Supervised Uncertainty Quantification for Segmentation with Multiple Annotations. arXiv [cs.LG] (2019). at <http://arxiv.org/abs/1907.01949>

20. Hoebel, K., Andrearczyk, V., Beers, A., Patel, J., Chang, K., Depeursinge, A., Müller, H. & Kalpathy-Cramer, J. An exploration of uncertainty information for segmentation quality assessment. in Medical Imaging 2020: Image Processing 11313, 381–390 (SPIE, 2020).

21. Kohl, S. A. A., Romera-Paredes, B., Meyer, C., De Fauw, J., Ledsam, J. R., Maier-Hein, K. H., Ali Eslami, S. M., Rezende, D. J. & Ronneberger, O. A Probabilistic U-Net for Segmentation of Ambiguous Images. arXiv [cs.CV] (2018). at <http://arxiv.org/abs/1806.05034>

22. Roy, A. G., Conjeti, S., Navab, N. & Wachinger, C. Inherent Brain Segmentation Quality Control from Fully ConvNet Monte Carlo Sampling. arXiv [cs.CV] (2018). at <http://arxiv.org/abs/1804.07046>

23. Carannante, G., Dera, D., Bouaynaya, N. C., Rasool, G. & Fathallah-Shaykh, H. M. Trustworthy Medical Segmentation with Uncertainty Estimation. arXiv [eess.IV] (2021). at <http://arxiv.org/abs/2111.05978>

24. Sagar. Uncertainty quantification using variational inference for biomedical image segmentation. Proceedings of the IEEE/CVF Winter Conference at <https://openaccess.thecvf.com/content/WACV2022W/VAQ/html/Sagar_Uncertainty_Quantification_Using_Variational_Inference_for_Biomedical_Image_Segmentation_WACVW_2022_paper.html>

25. Dohopolski, M., Chen, L., Sher, D. & Wang, J. Predicting lymph node metastasis in patients with oropharyngeal cancer by using a convolutional neural network with associated epistemic and aleatoric uncertainty. Phys. Med. Biol. 65, 225002 (2020).

26. Song, B., Sunny, S., Li, S., Gurushanth, K., Mendonca, P., Mukhia, N., Patrick, S., Gurudath, S., Raghavan, S., Tsusennaro, I., Leivon, S. T., Kolur, T., Shetty, V., Bushan, V. R., Ramesh, R., Peterson, T., Pillai, V., Wilder-Smith, P., Sigamani, A., Suresh, A., Kuriakose, M. A., Birur, P. & Liang, R. Bayesian deep learning for reliable oral cancer image classification. Biomed. Opt. Express 12, 6422–6430 (2021).

27. Dolezal, J. M., Srisuwananukorn, A., Karpeyev, D., Ramesh, S., Kochanny, S., Cody, B., Mansfield, A. S., Rakshit, S., Bansal, R., Bois, M. C., Bungum, A. O., Schulte, J. J., Vokes, E. E., Garassino, M. C., Husain, A. N. & Pearson, A. T. Uncertainty-informed deep learning models enable high-confidence predictions for digital histopathology. Nat. Commun. 13, 6572 (2022).

28. Dohopolski, M., Wang, K., Wang, B., Bai, T., Nguyen, D., Sher, D., Jiang, S. & Wang, J. Uncertainty estimations methods for a deep learning model to aid in clinical decision-making -- a clinician’s perspective. arXiv [cs.LG] (2022). at <http://arxiv.org/abs/2210.00589>

29. Nguyen, D., Sadeghnejad Barkousaraie, A., Bohara, G., Balagopal, A., McBeth, R., Lin, M.-H. & Jiang, S. A comparison of Monte Carlo dropout and bootstrap aggregation on the performance and uncertainty estimation in radiation therapy dose prediction with deep learning neural networks. Phys. Med. Biol. 66, 054002 (2021).

30. Tang, P., Yang, P., Nie, D., Wu, X., Zhou, J. & Wang, Y. Unified medical image segmentation by learning from uncertainty in an end-to-end manner. Knowledge-Based Systems 241, 108215 (2022).

31. Lei, W., Mei, H., Sun, Z., Ye, S., Gu, R., Wang, H., Huang, R., Zhang, S., Zhang, S. & Wang, G. Automatic segmentation of organs-at-risk from head-and-neck CT using separable convolutional neural network with hard-region-weighted loss. Neurocomputing 442, 184–199 (2021).

32. van Rooij, W., Verbakel, W. F., Slotman, B. J. & Dahele, M. Using Spatial Probability Maps to Highlight Potential Inaccuracies in Deep Learning-Based Contours: Facilitating Online Adaptive Radiation Therapy. Adv Radiat Oncol 6, 100658 (2021).

33. De Biase, A., Sijtsema, N. M., van Dijk, L., Langendijk, J. A. & van Ooijen, P. M. A. Deep learning aided oropharyngeal cancer segmentation with adaptive thresholding for predicted tumor probability in FDG PET and CT images. Phys. Med. Biol. (2023). doi:10.1088/1361-6560/acb9cf

34. Guo, C., Pleiss, G., Sun, Y. & Weinberger, K. Q. On Calibration of Modern Neural Networks. in Proceedings of the 34th International Conference on Machine Learning (eds. Precup, D. & Teh, Y. W.) 70, 1321–1330 (PMLR, 06--11 Aug 2017).

35. Izmailov, P., Vikram, S., Hoffman, M. D. & Wilson, A. G. G. What Are Bayesian Neural Network Posteriors Really Like? in Proceedings of the 38th International Conference on Machine Learning (eds. Meila, M. & Zhang, T.) 139, 4629–4640 (PMLR, 18--24 Jul 2021).

36. Jorge Cardoso, M., Li, W., Brown, R., Ma, N., Kerfoot, E., Wang, Y., Murrey, B., Myronenko, A., Zhao, C., Yang, D., Nath, V., He, Y., Xu, Z., Hatamizadeh, A., Myronenko, A., Zhu, W., Liu, Y., Zheng, M., Tang, Y., Yang, I., Zephyr, M., Hashemian, B., Alle, S., Darestani, M. Z., Budd, C., Modat, M., Vercauteren, T., Wang, G., Li, Y., Hu, Y., Fu, Y., Gorman, B., Johnson, H., Genereaux, B., Erdal, B. S., Gupta, V., Diaz-Pinto, A., Dourson, A., Maier-Hein, L., Jaeger, P. F., Baumgartner, M., Kalpathy-Cramer, J., Flores, M., Kirby, J., Cooper, L. A. D., Roth, H. R., Xu, D., Bericat, D., Floca, R., Kevin Zhou, S., Shuaib, H., Farahani, K., Maier-Hein, K. H., Aylward, S., Dogra, P., Ourselin, S. & Feng, A. MONAI: An open-source framework for deep learning in healthcare. arXiv [cs.LG] (2022). at <http://arxiv.org/abs/2211.02701>

37. Andrearczyk, V., Oreiller, V., Jreige, M., Vallières, M., Castelli, J., Elhalawani, H., Boughdad, S., Prior, J. O. & Depeursinge, A. Overview of the HECKTOR Challenge at MICCAI 2020: Automatic Head and Neck Tumor Segmentation in PET/CT. in Head and Neck Tumor Segmentation 1–21 (Springer International Publishing, 2021).

38. Gal, Y. & Ghahramani, Z. Dropout as a Bayesian Approximation: Representing Model Uncertainty in Deep Learning. in Proceedings of The 33rd International Conference on Machine Learning (eds. Balcan, M. F. & Weinberger, K. Q.) 48, 1050–1059 (PMLR, 20--22 Jun 2016).

39. Filos, A., Farquhar, S., Gomez, A. N., Rudner, T. G. J., Kenton, Z., Smith, L., Alizadeh, M., de Kroon, A. & Gal, Y. A Systematic Comparison of Bayesian Deep Learning Robustness in Diabetic Retinopathy Tasks. arXiv [stat.ML] (2019). at <http://arxiv.org/abs/1912.10481>

40. Abdar, M., Samami, M., Dehghani Mahmoodabad, S., Doan, T., Mazoure, B., Hashemifesharaki, R., Liu, L., Khosravi, A., Acharya, U. R., Makarenkov, V. & Nahavandi, S. Uncertainty quantification in skin cancer classification using three-way decision-based Bayesian deep learning. Comput. Biol. Med. 135, 104418 (2021).

41. Gal, Y. & Others. Uncertainty in deep learning. (2016).

42. McClure, P., Rho, N., Lee, J. A., Kaczmarzyk, J. R., Zheng, C. Y., Ghosh, S. S., Nielson, D. M., Thomas, A. G., Bandettini, P. & Pereira, F. Knowing What You Know in Brain Segmentation Using Bayesian Deep Neural Networks. Front. Neuroinform. 13, 67 (2019).

43. Roy, A. G., Conjeti, S., Navab, N. & Wachinger, C. Inherent Brain Segmentation Quality Control from Fully ConvNet Monte Carlo Sampling. in Medical Image Computing and Computer Assisted Intervention – MICCAI 2018 664–672 (Springer International Publishing, 2018).

44. Hoebel, K., Chang, K., Patel, J., Singh, P. & Kalpathy-Cramer, J. Give me (un)certainty -- An exploration of parameters that affect segmentation uncertainty. arXiv [eess.IV] (2019). at <http://arxiv.org/abs/1911.06357>

45. Mukhoti, J. & Gal, Y. Evaluating Bayesian Deep Learning Methods for Semantic Segmentation. arXiv [cs.CV] (2018). at <http://arxiv.org/abs/1811.12709>

46. Band, N., Rudner, T. G. J., Feng, Q., Filos, A., Nado, Z., Dusenberry, M. W., Jerfel, G., Tran, D. & Gal, Y. Benchmarking Bayesian Deep Learning on Diabetic Retinopathy Detection Tasks. arXiv [stat.ML] (2022). at <http://arxiv.org/abs/2211.12717>

47. Jaskari, J., Sahlsten, J., Damoulas, T., Knoblauch, J., Särkkä, S., Kärkkäinen, L., Hietala, K. & Kaski, K. K. Uncertainty-Aware Deep Learning Methods for Robust Diabetic Retinopathy Classification. IEEE Access 10, 76669–76681 (2022).

48. Taha, A. A. & Hanbury, A. Metrics for evaluating 3D medical image segmentation: analysis, selection, and tool. BMC Med. Imaging 15, 29 (2015).

49. Sherer, M. V., Lin, D., Elguindi, S., Duke, S., Tan, L.-T., Cacicedo, J., Dahele, M. & Gillespie, E. F. Metrics to evaluate the performance of auto-segmentation for radiation treatment planning: A critical review. Radiother. Oncol. 160, 185–191 (2021).

50. Andrearczyk, V., Oreiller, V., Vallières, M., Castelli, J., Elhalawani, H., Jreige, M., Boughdad, S., Prior, J. O. & Depeursinge, A. Automatic Segmentation of Head and Neck Tumors and Nodal Metastases in PET-CT scans. in Proceedings of the Third Conference on Medical Imaging with Deep Learning (eds. Arbel, T., Ayed, I. B., de Bruijne, M., Descoteaux, M., Lombaert, H. & Pal, C.) 121, 33–43 (PMLR, 2020).

51. Yang, J., Beadle, B. M., Garden, A. S., Schwartz, D. L. & Aristophanous, M. A multimodality segmentation framework for automatic target delineation in head and neck radiotherapy. Med. Phys. 42, 5310–5320 (2015).

52. Salzillo, T. C., Taku, N., Wahid, K. A., McDonald, B. A., Wang, J., van Dijk, L. V., Rigert, J. M., Mohamed, A. S. R., Wang, J., Lai, S. Y. & Fuller, C. D. Advances in Imaging for HPV-Related Oropharyngeal Cancer: Applications to Radiation Oncology. Semin. Radiat. Oncol. 31, 371–388 (2021).

53. Ren, J., Eriksen, J. G., Nijkamp, J. & Korreman, S. S. Comparing different CT, PET and MRI multi-modality image combinations for deep learning-based head and neck tumor segmentation. Acta Oncol. 60, 1399–1406 (2021).

